# The effect of functional calibration methods on gait kinematics in adolescents with idiopathic rotational deformity of the femur

**DOI:** 10.1101/2023.02.20.23286172

**Authors:** Ramon J. Boekesteijn, Myrthe P.F. van de Ven, Lise M. Wilders, Pepijn Bisseling, Brenda E. Groen, Katrijn Smulders

## Abstract

**Background:** Due to anatomical deviations, assumptions of the conventional calibration method for gait analysis may be violated in individuals with rotational deformities of the femur. We compared functional calibration methods with conventional methods in this group for 1) localization of the hip joint center (HJC) and knee axis orientation, and 2) gait kinematics.

**Methods:** Twenty-four adolescents with idiopathic rotational deformity of the femur underwent gait analysis and a CT scan. During standing, distance between HJCs and knee axis orientation were compared between calibration methods, with CT serving as reference for HJC estimation. Gait kinematics were compared using statistical parameter mapping (SPM).

**Findings:** Functional calibration methods estimated the HJC closer to the CT reference (26±21mm more lateral) than the conventional method (38±21mm more medial). In the full sample, orientation of the knee joint axis was not different between calibration methods, but in adolescents with increased femoral anteversion, the knee was more internally rotated with the functional method (3.3±6.2°). During gait, SPM revealed significantly more hip flexion, more internal hip rotation during the stance phase, less knee varus-valgus motion, and larger knee flexion angles when applying the functional calibration method.

**Interpretation:** Functional calibration methods better approximated the HJC, and showed a knee axis orientation that was more towards the direction of the deformity. This resulted in less knee joint angle crosstalk during gait. Although differences between calibration methods on gait kinematics were within clinically acceptable limits for the sagittal plane, relatively larger differences on transversal hip kinematics may hold clinical importance.

## 1. Introduction

Idiopathic rotational deformity of the femur in adolescents may cause internally or externally rotated gait, which can lead to pain complaints in the hip, knee, or ankle (1). Consequently, these adolescents can be limited in participating in daily-life activities (2), which can be a reason to visit a pediatric orthopedic surgeon. Although these complaints sometimes disappear spontaneously with growth over time, surgical management by a derotational osteotomy may be considered when pain complaints persist and conservative treatment fails.

To support clinical decision making regarding surgical procedures in adolescents with a rotational deformity of the femur, a combination of static and dynamic evaluations is optimal (3). The magnitude of a rotational deformity can be defined by femoral anteversion or retroversion, as measured on a computed tomography (CT) scan, or by physical examination. Interestingly, these evaluations pertain to static, unloaded situations – which poorly correlate with gait parameters (3) – while pain complaints often occur during dynamic activities. 3D gait analysis can likely provide relevant insights into aberrant lower extremity biomechanics and identify relevant compensations during walking, but these analyses have been limited to research settings (2-9).

A major limitation of 3D gait analysis in adolescents with rotational deformity of the femur is limited accuracy and reproducibility of kinematics in the transversal plane (10). The conventional method to compute 3D kinematics (i.e. Plug-in-Gait) relies on a number of anatomical assumptions and equations (11) which may, by definition, be violated in adolescents with rotational deformity of the femur. To determine the position of the hip joint centers (HJC), the conventional method uses equations with fixed relationships between marker positions on the pelvis, anthropometric measurements (i.e. leg length), and the hip joint center (HJC), which are derived from medical imaging studies in healthy adults (11). However, it is questionable whether these equations are also valid for adolescents (12), in particular for those with deviating anatomy. For the knee joint, rotational deformity of the femur may complicate defining the correct mediolateral knee axis. Errors made in the estimation of the HJC and orientation of the knee axis during calibration can propagate as offsets in hip rotation kinematics, and result in crosstalk of knee flexion towards knee varus-valgus motion during gait (12). Hence, alternative methods may be needed to improve 3D gait analysis in this population.

As an alternative for the conventional calibration method, functional calibration methods have been developed to be less dependent on marker placement and anthropometric measurements. With functional methods, the relative motion between two segments is the basis for a mathematically derived optimal location of the HJC and knee joint axis. For example, the symmetrical center of rotation estimation (SCoRe) (13) and symmetrical axes of rotation approach (SARA) (14) can be used to estimate the HJC and knee joint axis respectively. These methods assume the hip to be a ball-and socket joint with a fixed point of rotation, whereas the knee is modeled as a one degree of freedom hinge joint. In addition to a standing calibration, functional calibration trials are required to estimate the HJC and knee joint axis. These trials necessitate sufficient range of motion (RoM) of the knee and hip (15, 16), which has raised some concerns with respect to application of functional methods in clinical populations with restricted RoM, such as cerebral palsy (16). For adolescents with idiopathic rotational deformity of the femur, who do not have such limitations in active RoM, these functional calibration methods can potentially be an easily available option to improve gait analysis, but their performance compared to the conventional method still has to be evaluated.

In this study, we investigated the differences between conventional and functional calibration methods (i.e. SCoRe and SARA) in 1) localization of the HJC and orientation of the knee axis, and 2) gait kinematics in adolescents with rotational deformity of the femur. It was hypothesized that functional calibration methods would lead to a more accurate HJC estimation and a better definition of the knee joint axis, resulting in less crosstalk compared to conventional calibration methods.

## 2. Methods

### 2.1 Participants

Twenty-four adolescents (4 male, 20 female) with pain complaints due to a suspected idiopathic rotational deformity of the femur were included in this study. This sample was derived from a larger study evaluating 1) the value of gait analysis compared to CT and physical examination in adolescents with rotational deformity of the femur and/or tibia, and 2) the effects of a derotational osteotomy on pain and physical function. Patients were recruited from the outpatient pediatric orthopedic clinic of the Sint Maartenskliniek. Inclusion criteria were: 1) aged between 12-21 years old, 2) uni- or bilateral clinically deviating rotation of the hip in extension as measured with a goniometer (endorotation: male <25°or >65°, female <15°or > 60°; exorotation: <25°or >65°), 3) self-reported pain complaints in the leg related with rotational deviation. Patients were excluded from this study in case of presence of neuromuscular deficits impairing gait (incl. cerebral palsy), congenital malformation of the foot, mediolateral knee instability (i.e. > 15° varus or valgus RoM), deformity of the leg in the frontal plane as primary clinical problem, any other impairment causing gait or balance problems. The study was exempt from medical ethical review by the CMO Arnhem/Nijmegen (2019-5884), as it was not subject to the Medical Research Involving Human Subjects Act (WMO). All study procedures were conducted in accordance with the Declaration of Helsinki, and written informed consent was obtained from all participants, and their parents or guardians when required, prior to testing.

### 2.2 Gait analysis

Three-dimensional gait analysis was performed in the overground gait lab of the Sint Maartenskliniek Nijmegen. Kinematic data were collected using a ten camera motion capture system (*Vicon, Oxford, UK*) at a sample frequency of 100 Hz. Kinetic data was collected using two force plates (*Kistler Instruments, Hampshire, UK*; sample frequency = 1000 Hz) that were embedded in a 12m walkway. Passive reflective markers were attached to participants by an experienced lab technician. Marker placement on the anatomical landmarks was in accordance with the Vicon Lower Body marker set (11). For the standing calibration, additional markers were placed on the medial epicondyles and medial malleoli, in order to outline the anatomical knee and ankle axes. Thigh and tibia markers were subsequently moved anteriorly or posteriorly – if necessary – to align the Vicon knee and ankle axes with the anatomical transepicondylar and transmalleolar axes. For the functional calibration, additional markers were placed on the distal 1/3 of the femur and the proximal 1/3 of the tibia. Functional calibration trials consisted of hip RoM trials (i.e. active hip flexion, ab/adduction, extension, and circumduction (3 repetitions each)), and knee RoM trials (i.e. active knee lifts (5 repetitions)). Gait trials were collected after all calibration procedures. Participants were instructed to walk at a self-selected, comfortable speed. Five correct trials in which the subject hit each force plate with one foot were collected.

### 2.3 CT scan

Low-dose CT scans were made in a supine position according to the standard protocols of our hospital. Transversal slices (thickness = 3 mm) were made of the hip (caput femoris, collum, and trochanter), knee (femoral condyles and tibia plateau) and ankle (level of crural joint).

### 2.4 Data analysis

#### 2.4.1 Standing calibration and active range of motion trails

Three-dimensional marker data were processed in Vicon Nexus 2.9.2. Calibration trials were separately processed using 1) the conventional method, and 2) using functional methods as implemented in Vicon Nexus, including SCoRe and SARA along with optimal common shaping technique (17) (see Fig. 1 for a schematic representation). Subsequently, the distance between HJCs, and the knee axis orientation relative to the pelvis coordinative system were derived from the standing calibration trials. In order to verify that our participants achieved sufficient RoM during the functional calibration trials, RoM of the knee and hip were derived from the functional calibration trials.

**Fig. 1:**
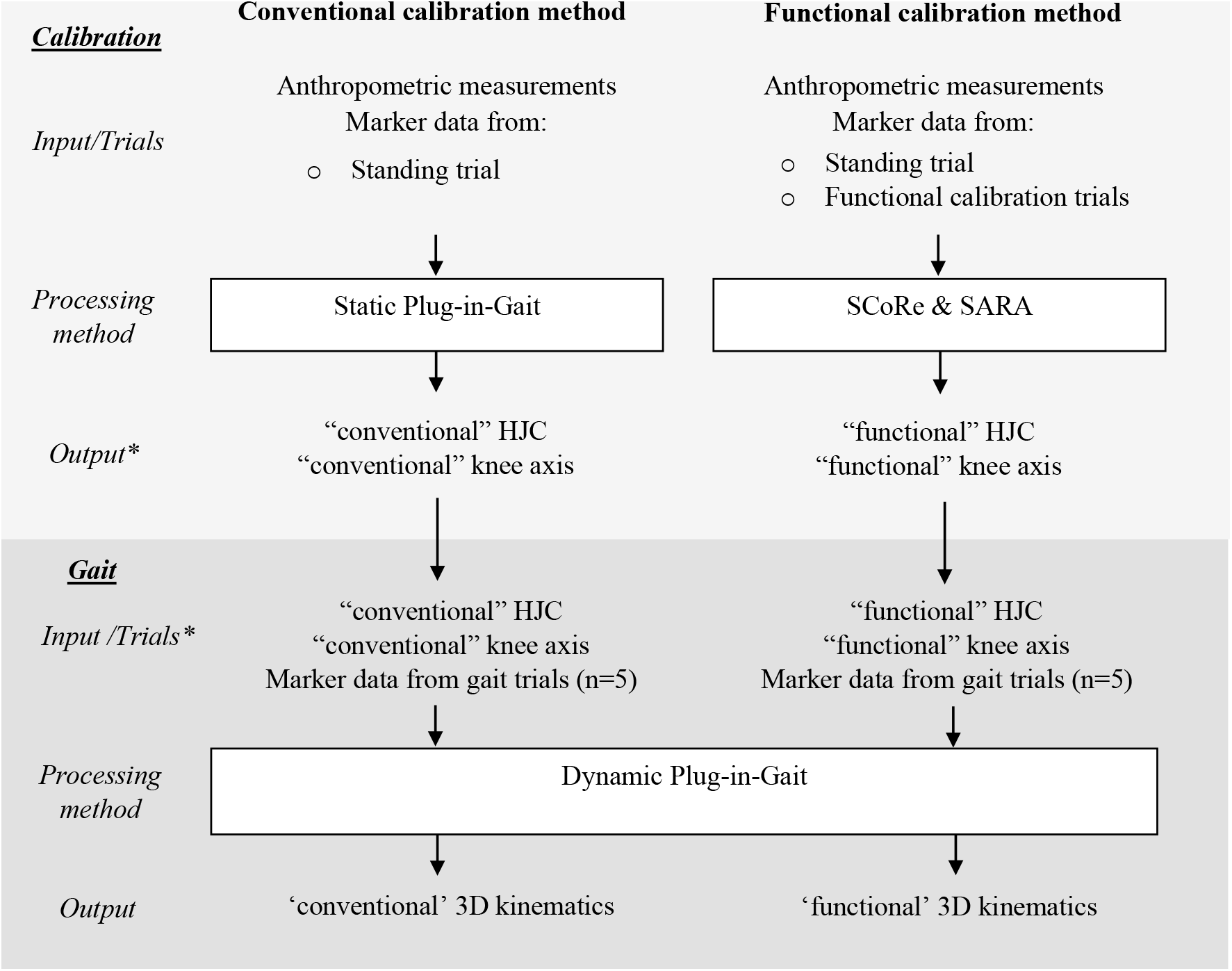
Simplified and schematic overview of the workflow for conventional and functional calibration methods. *For reasons of simplicity we only mentioned hip joint center (HJC) and knee axis as input/output, as these are the key differences between the two calibration methods.

#### 2.4.2 Gait

Marker data of the gait trials were interpolated, and data were filtered using a Woltring filter (MSE = 20). Gait events were detected by force plates and the position of the heel and toe markers, and were manually verified. Each gait trial was subsequently processed twice: using 1) input from the conventional calibration method, and 2) input from the functional calibration method (Fig. 1). Knee and hip kinematics were calculated, which were subsequently normalized to the duration of a stride. Only data of the most affected leg (i.e. the one with the most severe pain and/or radiological deformity) was included in the analysis. From the kinematics, the following discrete parameters were derived: 1) mean hip rotation during the stance phase, 2) knee varus-valgus RoM, and 3) maximum knee flexion. Data analysis was performed using custom written Matlab scripts.

#### 2.4.3 CT scan

Rotation of the femur (e.g. anteversion or retroversion) was defined as the angle between the line from the center of rotation of the caput femoris through the middle of the collum femoris, and a line along the posterior condyles of the distal femur, which was measured by a radiologist (AS) and an orthopedic surgeon (PB). If differences between raters were higher than 3°, inter-observer consensus was sought. For differences lower than 3°, the mean of both raters was taken. Subsequently, the distance between the center of rotation of the left and right caput femoris was calculated (RB, PB). If differences between raters were higher than 3 mm, inter-observer agreement was sought. For differences lower than 3 mm, the mean of both raters was taken.

#### 2.4.4 Statistical analysis

To estimate validity of the HJC localization, the distance between HJCs based on the CT scan (reference), those derived from the conventional calibration method, and functional calibration method were compared. A repeated ANOVA with post-hoc paired t-tests was used to compare these modalities. Differences in knee axis orientation during standing were compared between conventional and functional calibration methods using a paired t-test (α = 0.05).

Knee and hip kinematics were compared between conventional and functional calibration methods using one-dimensional statistical parameter mapping (SPM) (18) implemented in the SPM1D Matlab package. After testing for normality, a dependent sample t-test was conducted (SPM(t), α = 0.05). In addition, mean differences over the gait cycle with 95% and 99% CIs were plotted to obtain better insights in the effect sizes. Discrete parameters (i.e. mean hip rotation during the stance phase, maximum knee flexion, knee varus-valgus RoM) were compared between methods using a linear mixed model, with calibration method (functional vs. conventional) as fixed effect and participant ID as a random effect. Effects were reported as mean differences with 95% CI, with alpha set at 0.05. Statistical tests were conducted in RStudio 3.6.1. using the lme4 package.

As our sample was unbalanced in the distribution of the direction of rotational deformity of the femur (i.e. 22 adolescents with femoral anteversion versus 2 adolescents with retroversion), and the effects of the calibration method could potentially be opposite for these subgroups, we performed a post-hoc analysis, repeating all steps mentioned above, in which we removed the adolescents with increased femoral retroversion from the analysis.

## 3. Results

### 3.1 Patient characteristics and range of motion during functional calibrations

Mean age was 15 ± 2 years, height 1.68 ± 0.06 m, weight 57.6 ± 13.0 kg, and BMI 20.2 ± 3.8 kg/m^2^. Median femoral anteversion angle on CT was 34.6° (range: -26.7 – 63.2). Two adolescents presented with increased femoral retroversion (i.e. -26.7 and -6.8°), whereas the other twenty-two had increased femoral anteversion. During the functional calibration trials, active RoM of the hip was 63 ± 20° in the sagittal and 41 ± 13° in the frontal plane (Table 1). Furthermore, a RoM of 112 ± 17° was obtained for the knee in the sagittal plane during active knee lifts.

**Table 1:** Range of motion of the hip and knee during functional calibration trials.

| Parameter | Adolescents with rotational deformity of femur (n=24) |
| --- | --- |
| Hip flexion/extension RoM (°) | 63 (range: 34 – 130) |
| Hip ab/adduction RoM (°) | 41 (range: 24 – 87) |
| Knee flexion/extension RoM (°) | 112 (range: 68 – 143) |

### 3.2 Hip joint center localization and knee joint axis orientation

Estimated distance between the HJCs was significantly different between the three methods (F(2,46) = 120.8, p<0.001). HJC distances were 170 ± 13 mm using CT, 132 ± 20 mm using the conventional calibration methods, and 196 ± 21 mm using the functional method (i.e. SCoRe) (Fig. 2). Compared to CT, post-hoc comparisons indicated an underestimation of the HJC distance of 38 mm (95% CI: 30, 46; p<0.001) in the conventional calibration method, whereas the functional calibration method overestimated HJC distance with 26 mm (95% CI: 18, 34; p<0.001). Post-hoc removal of the participants with femoral retroversion did not yield different results (Supplementary File 1 – Table 1).

**Fig. 2:**
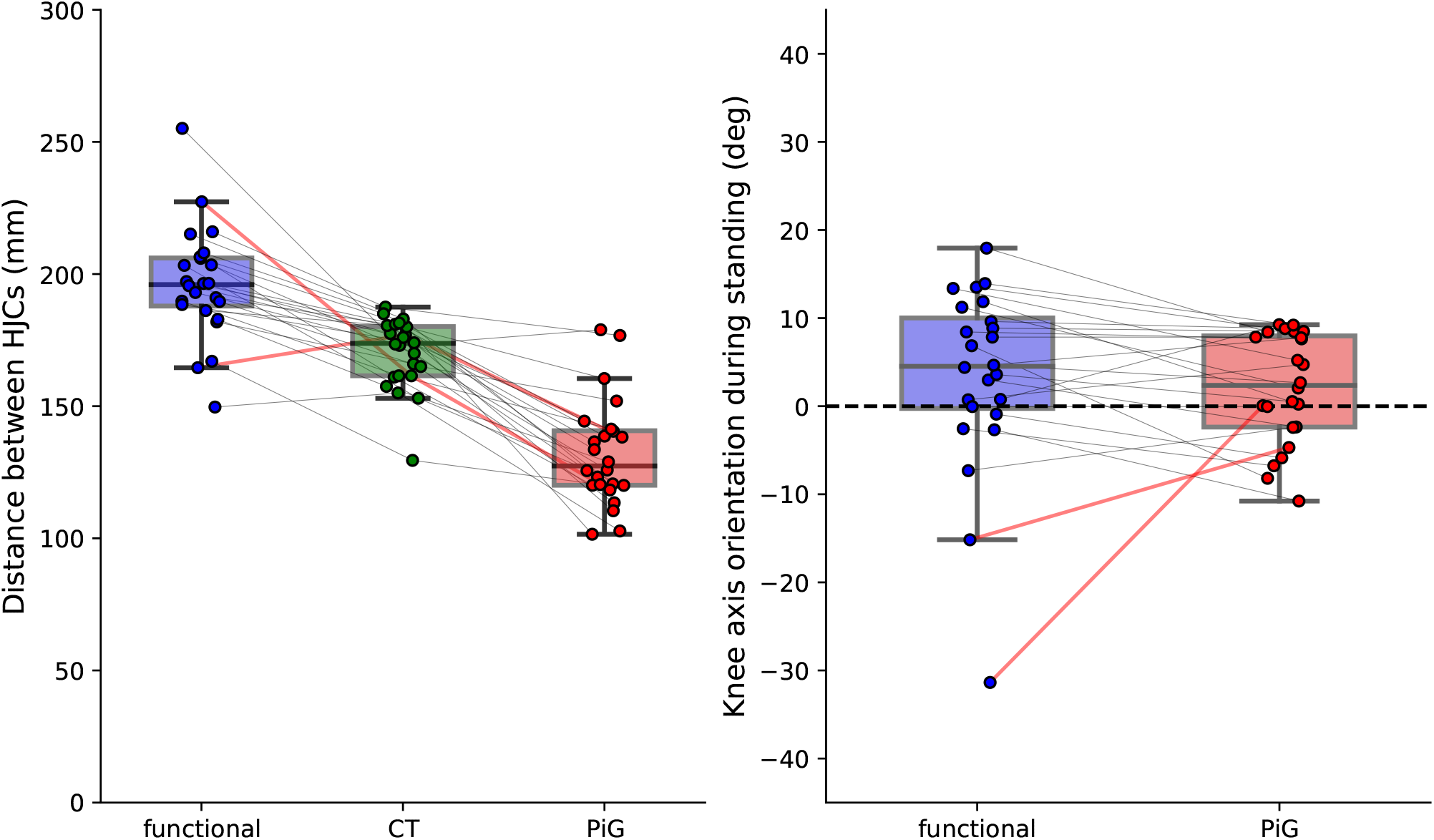
Differences in hip joint center (HJC) localization and orientation of the knee axis during the static, standing calibration. Boxes represent the interquartile range, while the whiskers indicate the upper and lower quartiles. Individual data is showed by scatter overlay, with all measures within a participant connected through lines. The two participants with increased femoral retroversion are highlighted in red.

Orientation of the knee axis during standing was not significantly different between the functional and conventional calibration method (mean difference = 1.3 deg, 95% CI: -2.7, 5.2 ; p = 0.513). In both methods, knee axis orientation was directed internally in the majority of participants (Fig. 2). However, removal of the adolescents with retroversion led to a significant difference between the calibration methods on knee axis orientation (t(21) = 2.684, p = 0.014)). In the subgroup with only the adolescents with increased femoral anteversion, the functional calibration method resulted in a knee axis orientation which was 3.3° (95% CI: 0.7, 5.8) more internally rotated compared to the conventional calibration method (Supplementary File 1 - Table 1).

### 3.3 Effect of calibration method on gait kinematics

In the full sample of adolescents with a rotational deformity of the femur, functional calibration methods yielded significantly different hip kinematics compared to the conventional calibration method, as illustrated by SPM analysis (Fig. 3). SPM revealed that the hip was flexed more between 0 – 43% and 53 – 100% of the gait cycle using the functional calibration method compared to the conventional method. Over the whole gait cycle, the hip was approximately 4-5° more in adduction with the functional calibration method compared to the conventional method. In the transversal plane, the functional calibration method showed 5-10 degrees more external rotation during the swing phase (61% – 89%). Mean hip rotation during the stance phase was not significantly different between calibration methods (mean diff = 0.6 deg, 95% CI: -0.7, 1.9; p = 0.367)

**Fig. 3:**
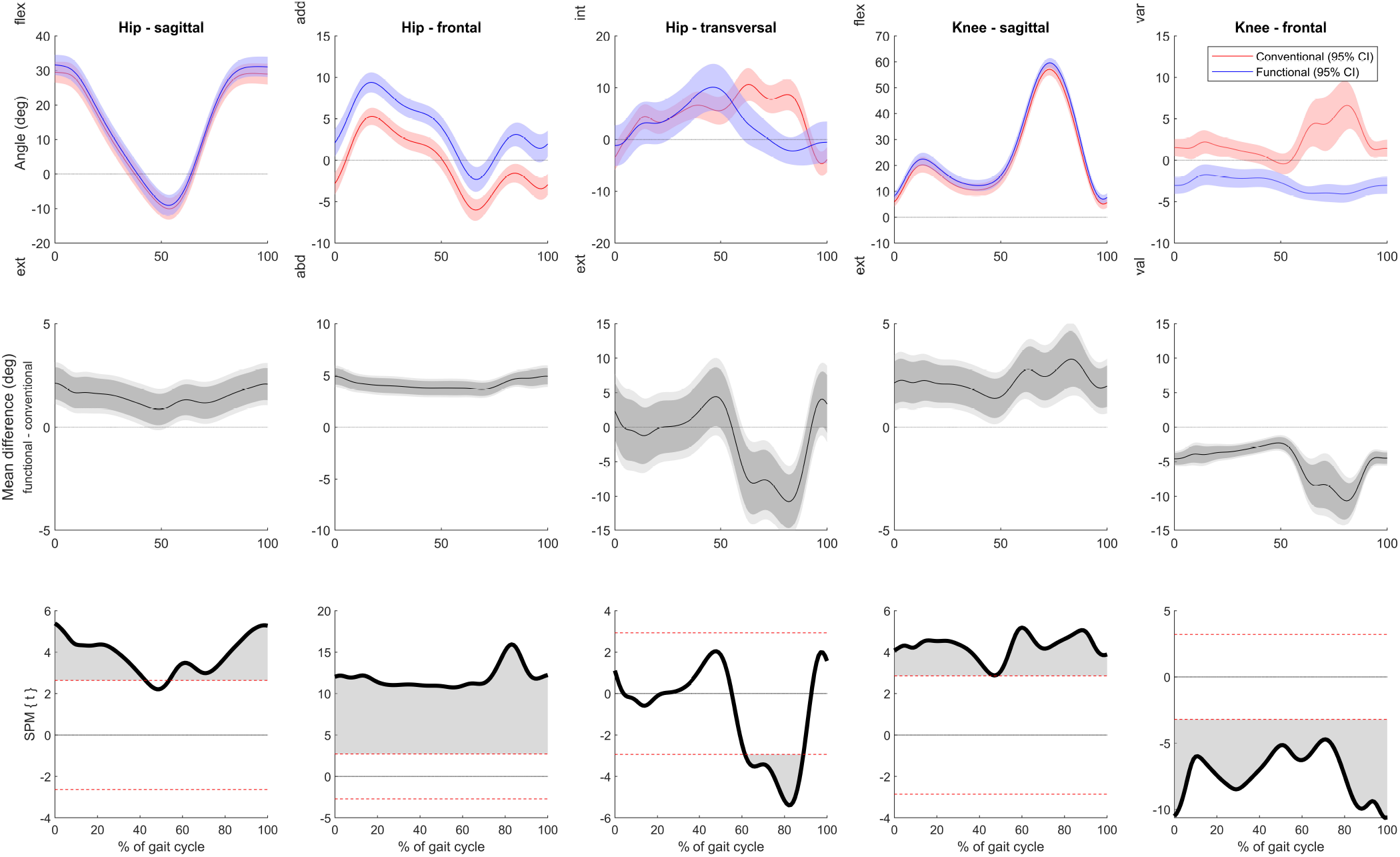
Hip and knee kinematics for the functional (blue) and conventional calibration method (red). Group means and 95% confidence intervals are shown in the top row. The middle row shows the mean differences with the 95% (dark grey) and 99% confidence interval (light grey). Results of statistical parameter mapping (SPM) are displayed in the bottom row. Significant differences between the models are denotated by the grey marked area.

Considering the knee joint, SPM revealed that the knee was more flexed over the whole gait cycle (0-100%) using the functional calibration method (Fig. 3). Maximum knee flexion was 2.4 deg (95 % CI: 1.8, 3.0; p<0.001) higher with the functional calibration method compared to the conventional method. Finally, frontal plane knee kinematics were significantly different between the two calibration methods over the whole gait cycle (0-100%), with the functional calibration method placing the knee in valgus and the conventional method placing the knee in varus. Total knee varus-valgus RoM was 6.0 deg (95% CI: 5.3, 6.7; p<0.001) lower using functional calibration methods compared to conventional calibration methods.

Post-hoc removal of adolescents with femoral retroversion from the analysis changed our results on the effect of the calibration method on hip and knee kinematics (Supplementary File 1 – Fig. 1). First, in adolescents with femoral anteversion, SPM indicated that the functional calibration method yielded significantly higher internal rotation angles during terminal stance (41 – 52%) compared to the conventional calibration method. Mean hip rotation during stance in this subset was 2.5° (95% CI: 1.4, 3.5; p<0.001) more internal in the functional compared to the conventional calibration method, whereas this was not significantly different in the full sample. As shown in Fig. 4, adolescents with increased femoral retroversion had more external hip rotation with the functional calibration method compared to the conventional calibration method, which was opposite to the mean effect in adolescents with increased femoral anteversion. Second, SPM results of the sagittal hip and knee kinematics were slightly different in post-hoc analysis compared to analysis of the full sample. In the adolescents with increased femoral anteversion, the hip was flexed more between 0-37%, 58-66%, and 75-100% of the gait cycle when using the functional calibration method compared to conventional calibration method. For the knee, larger flexion angles were obtained between 0-42 % and 51-100% with the functional calibration method compared to the conventional calibration method. Although SPM results of sagittal these knee and hip kinematics were different in the post-hoc analysis, the mean differences remained relatively similar compared to analysis of the full sample (Supplementary File – Fig. 1).

**Fig. 4:**
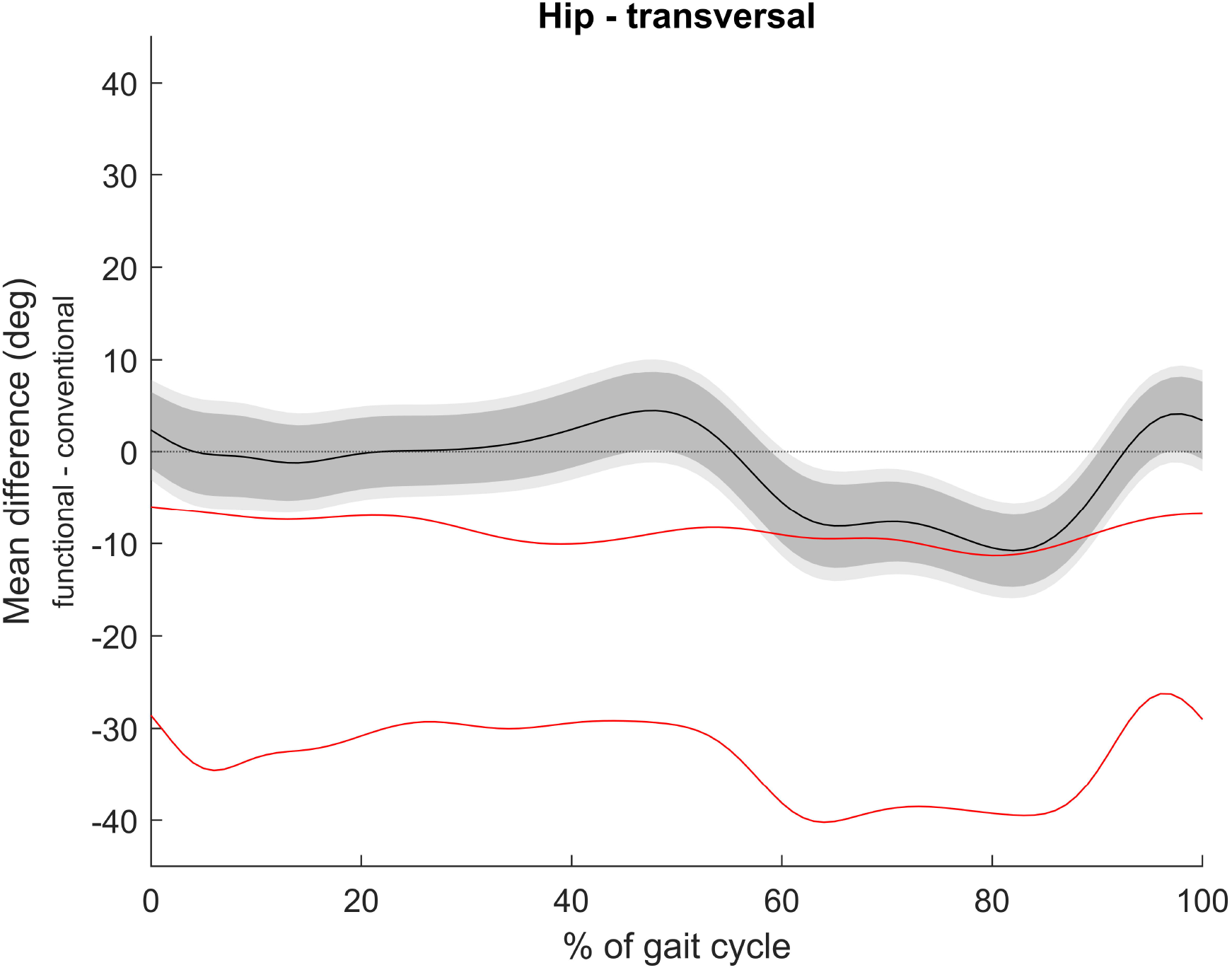
Mean differences between the two calibration methods (i.e. functional – conventional) for hip internal/external rotation. Data for the complete group is showed as mean with 95% (dark grey area) and 99% (light grey area). Individual data of the two adolescents with increased femoral retroversion is highlighted in red.

## 4. Discussion

Application of functional calibration methods resulted in more lateral estimation of the HJC location compared to the CT reference, whereas the conventional calibration method positioned the HJC more medially. Only for adolescents with increased femoral anteversion, but not in those with increased femoral retroversion, the knee axis was more internally rotated with functional methods compared to the conventional calibration methods. During gait, functional calibration methods resulted in less knee varus-valgus RoM, and larger knee and hip flexion angles compared to the conventional method. Finally, the hip was more abducted and more externally rotated during the swing phase using the functional calibration methods compared to the conventional method.

### 4.1 Localization of hip joint centers and orientation of knee joint axis

Our finding that the conventional calibration method and functional calibration methods (i.e. SCoRe) were not fully accurate in determining the HJC’s is in line with the literature. Multiple studies found that the Davis predictive method (11), which is used in the conventional calibration method, places the HJC approximately 10-15 mm more medial compared to a reference HJC as obtained with dual fluoroscopy (19), 3D ultrasound (20), low-dose X-rays (21), and CT scans (22). For functional calibration methods, three studies reported that SCoRe placed the HJC ∼5-10 mm more lateral compared to the reference HJC (19-21). Our findings are in agreement with these studies, although the deviation from the reference was larger in our study (i.e. 19 mm more medial in the conventional calibration method, and 13 mm more lateral with functional calibration methods). In contrast, Assi *et al*. did not find significant differences in HJC localization in the mediolateral direction when conventional methods and SCoRe were compared to low-dose X-ray reference (23).

We found no differences between the conventional calibration method and functional calibration methods (i.e. SARA) in the orientation of the knee axis during the standing calibration trial, which is also in agreement with previous studies (9, 24, 25). However, post-hoc analysis indicated that the effect of the calibration method on knee axis orientation may be dependent on the direction of the rotational deformity. Our results imply that a functional calibration method including SARA accommodated for more extreme cases of internal and external orientation of the knee axis, whereas in the conventional calibration method the knee axis orientation was oriented in a relatively neutral position between -10 and 10 degrees. Importantly, Sauret *et al*. and Passmore *et al*. showed that both functional models (i.e. SARA) and the conventional calibration method deviated on average 5-10 degrees (external) from the actual transepicondylar axis obtained using biplanar radiographs (24) and 3D ultrasound measurement (25). Taken together, this suggests that function calibration methods might lead to more accurate estimation of knee axis orientation in adolescents with rotational deformity of the femur, but that both the knee axis derived from functional and conventional calibration methods could still differ from the actual transepicondylar axis.

### 4.2 Effect of calibration method on gait kinematics

During gait, we observed differences in hip and knee kinematics between the functional and conventional calibration method. A part of these kinematic differences can be the direct consequence of differences in HJC localization between the two methods. Using trigonometry, lateral translation of the HJC of ∼3 cm compared to the pelvis midline would indeed result in the offset in frontal plane hip kinematics of ∼4° towards adduction that we found, when assuming a femoral length of 25% of a participants’ height (26). Similarly, lateral translation of the HJC will place the knee joint towards valgus alignment. This may thus explain the shift in kinematics from knee varus to valgus when using the functional calibration methods.

The functional calibration methods resulted in lower knee varus-valgus RoM accompanied by a larger maximum knee flexion angle compared to conventional calibration methods, and therefore seemed to reduce crosstalk. Crosstalk is defined as the incorrect measurement of knee flexion as knee varus-valgus movement, due to a suboptimal approximation of the mediolateral knee axis (27-29). In part, this error may arise from improper placement of the knee and thigh marker when using conventional calibration methods. When comparing our results with other literature, knee varus-valgus RoM during gait derived from the functional calibration method was found to be close to fluoroscopy findings in healthy adults, which were 3.4° during a 60 degree flexion task (30), and 5.4° (stance) and 6.3° (swing) during walking (31). In our data, a direct result of this lower cross-talk was that higher knee flexion angles were obtained with functional calibration methods compared to the conventional calibration method, although the magnitude of this difference (i.e. 2.4°) can be considered to be marginal. Similarly, the effect of the calibration method on hip flexion angles was small. Hence, this again strengthens the notion that these sagittal plane gait parameters are robust outcomes of gait analysis with clinically acceptable errors (10).

In line with our findings on knee joint axis orientation, the calibration method had a significant effect on hip rotation kinematics. Functional calibration methods yielded a different kinematic profile in the transversal plane compared to the conventional calibration method, characterized by more external rotation during swing and 2.5° more internal rotation during stance in the adolescents with femoral anteversion only. Effects of the calibration method on hip rotation kinematics seemed to be opposite in adolescents with increased femoral retroversion, although a larger sample would be needed to further generalize this. Our findings are not in agreement with the study of Passmore *et al*., who did not find clear differences in hip kinematics between a conventional and functional calibration method (i.e. SARA) (9). This may be explained by the fact that they included a different patient group (incl. adolescents with cerebral palsy) and only used SARA in their functional calibration method. Noteworthy is that in their study, the functional calibration method as well as the conventional calibration method showed offsets from transversal hip kinematics obtained using low-dose X-rays (i.e. both were more external than the reference). This could only partially be explained by a different knee axis definition (i.e. transepicondylar axis vs. condylar knee axis). Clinicians and researchers should thus be careful with interpretation of hip rotation kinematics, in particular as indicator for derotational osteotomy (32, 33), as the effect of the calibration method as well as previous reported measurement error for this parameter (10) may approach values that seem relevant for clinical decision making (33).

This study had a number of limitations. First, we had no ‘ground truth’ (i.e. fluoroscopy) available to validate the gait kinematics in this study, and thus cannot directly interpret our results in terms of superiority of one of the two methods with regard to gait kinematics. Secondly, only two patients with femoral retroversion were included in the sample, which limited opportunities for subgroup analysis. This may be worth a future investigation based on our post-hoc analysis. Thirdly, not all adolescents reached the recommended level of RoM for functional calibration trials (i.e. 60° hip flexion, 30° hip ab/adduction (15)). This may have resulted in inaccurate estimations of the HJC and knee joint axis using the functional method, limiting its potential advantage. Finally, the test-retest reliability of the functional method was beyond the scope of this study, but is an important aspect to consider when deciding which calibration method provides better results. Currently, there are some indications in typically developing children that reliability of gait kinematics may improve by using functional calibration methods, but compelling evidence is lacking (34).

## 5. Conclusions

Functional calibration methods resulted in closer approximation of the anatomical HJC and potentially a better orientation of the knee joint axis compared to conventional calibration methods, resulting in less knee joint angle crosstalk during gait. Effects of the calibration method on sagittal knee and hip kinematics were within clinically acceptable limits. However, relatively larger differences between calibration methods in transversal plane hip kinematics may hold clinical relevance, particularly for a population with deformities in this specific plane. Hence, cautious interpretation of this outcome is warranted for adolescents with rotational deformity of the femur.

## Data Availability

All data produced in the present study are available upon reasonable request to the authors

## Acknowledgements

We want to thank Arjan Steenbakkers (AS) for their contribution to analysis of the radiographic images, and Lisa van der Wiel for her assistance during the data collection.

---

**Supplementary File 1 – post-hoc analysis: removal of 2 adolescents with increased femoral retroversion**

**Table 1:** Post-hoc analysis on the effects of calibration method on HJC localization and knee axis orientation.

|  | CT | Functional calibration | Conventional calibration | Statistical tests |
| --- | --- | --- | --- | --- |
| **Full sample (n=24)** |  |  |  |  |
| Distance between HJCs (mm) | 170 (13) | 196 (21) | 132 (20) | F(2,46) = 120.8, p<0.001 |
| Knee axis orientation (deg) | - | 3.4 (10.6) | 2.1 (6.2) | t(23) = -0.664, p = 0.513<br>mean diff = -1.3 deg (-5.2, 2.7) |
| **Post-hoc analysis (n=22)** |  |  |  |  |
| Distance between HJCs (mm) | 170 (14) | 196 (20) | 133 (21) | F(2,42) = 130.0, p<0.001 |
| Knee axis orientation (deg) | - | 5.8 (6.5) | 2.5 (6.3) | t(21) = 2.684, p = 0.014;<br>mean diff = 3.3 deg (0.7, 5.8) |

**Fig. 1:**
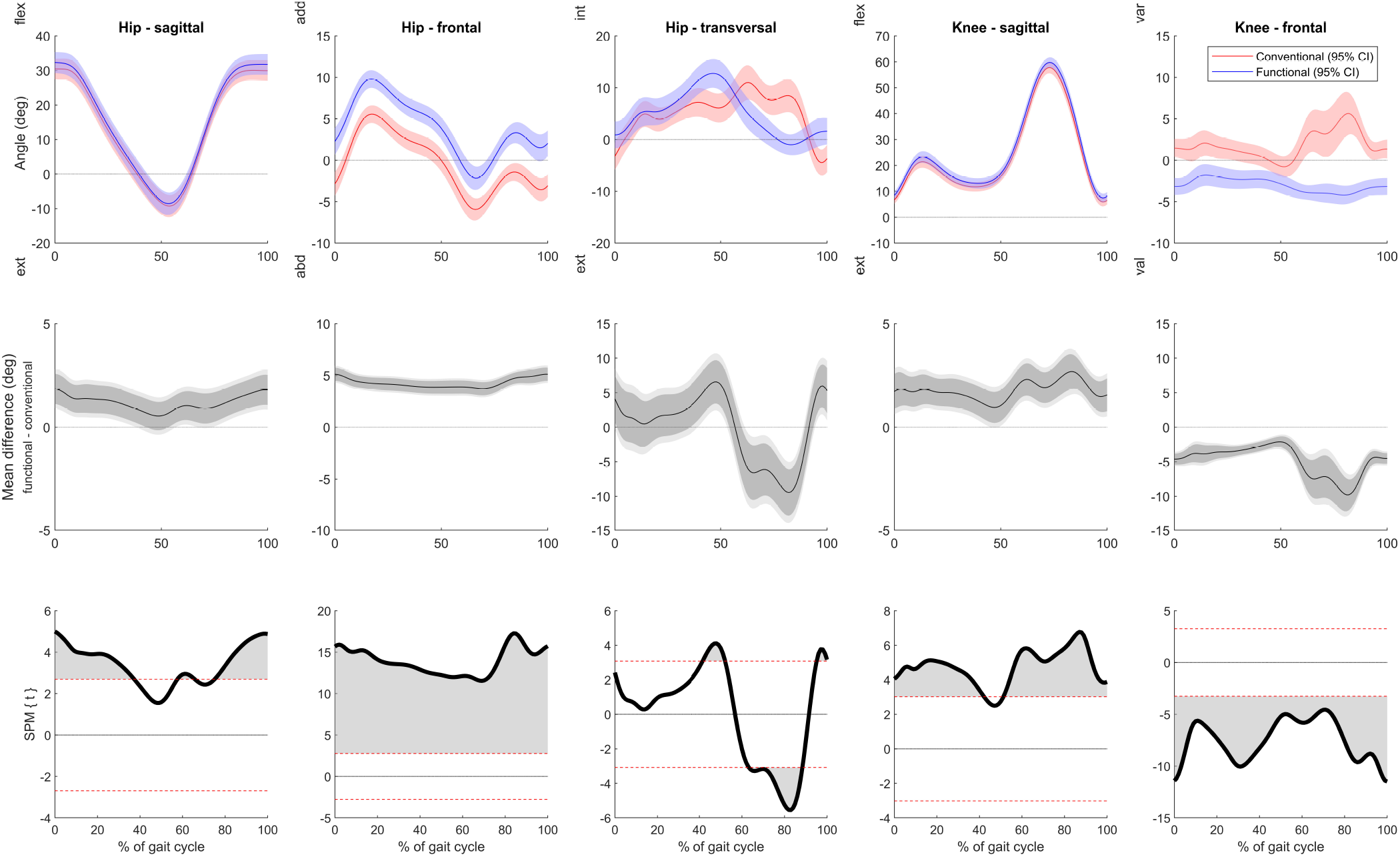
Hip and knee kinematics for the functional (blue) and conventional method (red) after removal of 2 adolescents with increased femoral retroversion. Group means and 95% confidence intervals are shown in the top row. The middle row shows the mean differences with the 95% (dark grey) and 99% confidence interval (light grey). Results of statistical parameter mapping (SPM) are displayed in the bottom row. Significant differences between the models are denotated by the grey marked area.

